# Wearable technology to capture arm use of stroke survivors in home and community settings: feasibility and early insights on motor performance

**DOI:** 10.1101/2023.01.25.23284790

**Authors:** Marika Demers, Lauri Bishop, Amelia Cain, Joseph Saba, Justin Rowe, Daniel Zondervan, Carolee Winstein

**Affiliations:** School of Rehabilitation, Université de Montréal, Montreal (Qc), Canada; Division of Biokinesiology and Physical Therapy, Herman Ostrow School of Dentistry, University of Southern California, Los Angeles, CA, USA; Flint Rehabilitation Devices, Irvine (CA), USA; Department of Neurology, Keck School of Medicine, University of Southern California, Los Angeles, CA

**Keywords:** upper extremity, rehabilitation, stroke, wearable electronic devices, motor skills, motor activity, self-efficacy

## Abstract

**Objective:** To establish short-term feasibility and usability of wrist-worn wearable sensors to capture arm/hand activity of stroke survivors and to explore **the association between** factors related to use of the paretic arm/hand.

**Methods:** 30 chronic stroke survivors were monitored with wrist-worn wearable sensors during 12h/day for a 7-day period. Participants also completed standardized assessments to capture stroke severity, arm motor impairments, self-perceived arm use and self-efficacy. Usability of the wearable sensors was assessed using the adapted System Usability Scale and an exit interview. Associations between motor performance and capacity (arm/hand impairments and activity limitations) were assessed using Spearman’s correlations.

**Results:** Minimal technical issues or lack of adherence to the wearing schedule occurred, with 87.6% of days procuring valid data from both sensors. Average sensor wear time was 12.6 (standard deviation: 0.2) h/day. Three participants experienced discomfort with one of the wristbands and three other participants had unrelated adverse events. There were positive self-reported usability scores (mean: 85.4/100) and high user satisfaction. Significant correlations were observed for measures of motor capacity and self-efficacy with paretic arm use in the home and the community (Spearman’s correlation ρs: 0.44-0.71).

**Conclusions:** This work demonstrates the feasibility and usability of a consumer-grade wearable sensor to capture paretic arm activity outside the laboratory. It provides early insight into stroke survivors’ everyday arm use and related factors such as motor capacity and self-efficacy.

**Impact:** The integration of wearable technologies into clinical practice offers new possibilities to complement in-person clinical assessments and to better understand how each person is moving outside of therapy and throughout the recovery and reintegration phase. Insights gained from monitoring stroke survivors arm/hand use in the home and community is the first step towards informing future research with an emphasis on causal mechanisms with clinical relevance.

## 1. Background

Arm impairments are highly prevalent after stroke (up to 80%)^1^ and can impact one’s ability to complete activities of daily living and consequently contribute to reductions in community participation and quality of life.^2,3^ The transfer of skills gained in the therapy context to real-life situations remains a challenge for stroke rehabilitation. Consistent with the terminology of the International Classification of Functioning,^4^ the activity domain can be separated into the capacity for activity (i.e., what one can do; assessed by standardized tests in structured settings) versus performance in daily activities (i.e. what one does in the home/community). For example, persistent difficulty with hand movements or reduced hand use in daily activities were reported in 71% of stroke survivors with full arm motor function recovery measured by the Fugl-Meyer Assessment.^5^ Similarly, a recent longitudinal study showed that 59% of stroke survivors that received outpatient rehabilitation care improved capacity for activity but not performance in daily activities.^6^ This disparity highlights the need to capture arm use outside clinical settings to gain more insights into real-life motor performance.

Knowledge of spontaneous motor performance can help clinicians plan effective and efficient rehabilitation interventions, thereby maximizing the potential for lasting functional gains.^7^ However, clinical assessments are traditionally done in highly-structured environments, which may not accurately reflect the behaviors exhibited in the natural environment with all its unpredictable and changing characteristics.^8,9^ Wearable technology offers new means of data acquisition in ecologically relevant environments to potentially inform stroke rehabilitation.^10^ It can also be used as an intervention modality to encourage health-promoting behaviors that may reduce disability in stroke survivors.^11^ Longitudinal monitoring and promotion of paretic arm/hand use informed through wearable technology may be key to encourage a virtuous cycle of arm use and promote self-rehabilitation in the community.^12^ Previous work demonstrated that research-grade wrist-worn wearable sensors are a valid and reliable means to capture arm/hand use in stroke survivors^13,14^ and to distinguish between paretic and less-affected arms.^15^ However, research-grade sensors have many limitations for clinical use: data needs to be processed offline, the key output, i.e., activity count, may be difficult to interpret, and most systems are expensive and not user-friendly.^11,16,17^ Requirements such as user-friendliness, robustness, and ability for online data processing are crucial for clinical adoption.^18^ Currently, there is a plethora of commercially available consumer-grade devices to capture physical activity behavior, yet, limited options exist to accurately capture arm/hand movement of stroke survivor with a wide range of motor impairments.^11,17,19^ The consumer-grade MiGo system (Flint Rehabilitation Devices, Irvine, CA) was developed to monitor the activity (i.e., arm/hand use and mobility) of stroke survivors in their homes and communities, and to address some of the limitations of existing technologies. Our previous work supported the accuracy of MiGo to capture time in active movement for each arm in a laboratory setting in chronic stroke survivors.^20^ However, feasibility and usability in the natural setting is necessary to **ultimately** translate the use of wearable technologies into meaningful therapeutic tools.

Wearable technology in combination with clinical measures provides valuable data to better understand how each person is moving outside of therapy and throughout the recovery and reintegration phase. We must examine not only a stroke survivors’ paretic arm use which can be captured through wearable technology, but also how these data supplement and relate to lab- or clinic-based assessments. Ultimately, with future research, this combination of data may help us better understand why improvements in motor capacity in some people do not necessarily translate to better performance in daily activities, while in others they do. This understanding has the potential to provide valuable insights for the development of new treatments to promote functional recovery.^21^

This project aims to 1) establish short-term feasibility and usability of wrist-worn wearable sensors to capture arm/hand movement behavior in the unsupervised home/community environment, in chronic stroke survivors, and 2) demonstrate the clinical relevance of wearable sensors through exploring the associations between paretic arm/hand use and both motor capacity and self-efficacy in the natural environment. For objective 1, feasibility is assessed using four metrics: 1) adherence, 2) technical issues and malfunction, 3) safety and comfort, and 4) acceptance and satisfaction. Our milestones are to achieve no severe adverse events, >80.0% of days with valid data collected from both sensors, positive self-reported usability scores (>70.0%)^22^ and user’s satisfaction. For objective 2, we hypothesize that paretic arm use (i.e., motor performance) will be **associated with** measures of motor capacity and self-efficacy. We expect that self-reported measures of arm use may have a weaker relationship with sensor-based measures of motor performance, as self-reported measures are known to vary greatly with direct measures of activity.^23–25^

## 2. Methods

### 2.1. Design

This study used an observational study design (as part of a larger study) to examine both the feasibility of wearable sensors to capture arm/hand use and locomotor behavior and also the feedback preferences of stroke survivors. Only the feasibility for monitoring arm/hand use is reported here.

### 2.2. Participants

Participants were included if they had an ischemic or hemorrhagic stroke, were aged >18 years old, lived at home and were able to communicate in English. Exclusion criteria were: unilateral spatial neglect (positive score on 2/3 screening measures^26^), assistance for ambulation, severe cognitive or language impairments, or other medical condition that can interfere with participation. We purposefully recruited participants with a broad range of motor impairments and included participants with mild-moderate aphasia to better generalize our results. We initially recruited participants who took part in our previous validity study^20^ and recruited additional participants using the IRB-approved Registry for Healthy Aging Database to reach our *a priori* target sample size of 30 participants. All participants were fully informed of the procedures involved and provided informed consent. The study complies with the Declaration of Helsinki. Study procedures were approved by the IRB at the University of Southern California (HS 20-00015).

### 2.3. Wearable sensor

Participants wore the MiGo activity watch on each wrist to capture arm movements. MiGo is a six degrees of freedom inertial measurement unit equipped with an adjustable silicone wristband and a Bluetooth radio. Accelerometer data were analyzed using a custom built-in active time counter algorithm.^16^ Each watch logged their respective data to a persistent block of flash memory. While MiGo has a screen that can display feedback metrics, this information was disabled to remove the bias of providing feedback. Participants were sent home with a cellular gateway (Tenovi Health, Irvine, CA). Every three hours, the gateway scanned for the sensors, connected to them, read, and relayed their logs to a HIPAA compliant server.

### 2.4. Procedures

Participants took part in two in-lab visits and were monitored at home for seven days. During Visit 1, participants completed a battery of standardized assessments. Information on how to wear and charge the sensors were provided. Participants were instructed to wear the sensors for 12h each day, continue their typical activities, and charge the sensors each night. Since the activity watches are not waterproof, we asked participants to remove the sensors for showering, bathing, or swimming. For the less-affected arm, the silicone band was replaced by an elastic band to facilitate donning/doffing. A power supply was provided, and each participant was given a ‘Tips’ sheet with reminders for daily wear, sensor care, and precautions (see Supplementary material). The research team connected daily to a remote monitoring website to monitor adherence and identify any system malfunction. Participants were contacted after an initial 48hs and/or when data were missing to resolve technical issues or answer questions.

Equipment was returned during Visit 2. Participants completed three surveys on usability, self-efficacy, and perceived arm/hand use. Afterwards, a summary of their motor performance over the monitoring period was offered. Experience with the sensors, adverse events and technical issues reported were captured using a semi-structured interview (audio-recorded). The interview followed a detailed interview guide (see Supplementary material) about experience with the wearable sensors, feedback preferences and factors influencing behavior and recovery.

### 2.5. Outcome measures

#### 2.5.1. Clinical measures

Standardized assessments were used to characterize cognitive function (Montreal Cognitive Assessment),^27^ stroke severity (National Institutes of Health Stroke Scale),^28^ and handedness (Edinburgh Handedness Inventory).^29^ At Visit 1, the upper extremity Fugl-Meyer Assessment (FMA-UE),^30^ the Chedoke Arm and Hand Activity Inventory-7 (CAHAI),^31^ and the Rating of Everyday Arm-use in the Community and Home^32^ were administered to capture arm/hand motor impairments, activity limitations and perceived use, respectively. At Visit 2, the adapted Systems Usability Scale (SUS; usability),^33^ the Motor Activity Log-14 (self-perceived arm/hand function)^34^ and the Confidence in Arm and Hand Movement (self-efficacy)^35^ were collected. Data were entered in the REDCap platform (Vanderbilt University, Nashville, TN) and verified by another member of the research team.

The FMA-UE assesses reflex action, movement and coordination of the shoulder, elbow, forearm, wrist and hand.^30^ Each item is scored by visual observation on a three-point ordinal scale (0=cannot perform; 1=performs partially; 2=performs fully). Each item is added for a maximal score of 66 indicating complete motor recovery.

The CAHAI is a performance-based assessment of arm/hand functional recovery. It comprises seven bimanual tasks.^31^ Each task is scored on a 7-point scale (1=total assistance; 7=total independence). Higher scores indicate greater functional independence.

The Confidence in Arm and Hand Movement is a self-reported measure of self-efficacy for paretic arm/hand function in social or home/community contexts.^35^ It consists of 20 questions scored on a visual analogue scale (0=very uncertain, 100=very certain). The scores are averaged to provide a total scale score between 0 to 100, with higher scores showing greater self-efficacy.

The Rating of Everyday Arm-use in the Community and Home is a self-reported measure of paretic arm use outside the clinical setting.^32^ It comprises two scales, based on whether the dominant or non-dominant arm is affected. Each scale consists of 6 categories of use (0=No Use; 5=Full Use).

The Motor Activity Log is a 14-item self-reported measure administered by semi-structured interview.^34^ The shorter version was selected over the original Motor Activity Log to minimize administration burden of multiple surveys. The psychometric properties are similar to the original Motor Activity Log.^34,36^ Participants are asked to determine (a) how much (amount of use scale), and (b) how well (quality of movement scale) they used the paretic arm/hand in the past week. Scoring ranges from 0 (never use the paretic arm/hand) to 5 (same as pre-stroke). For each scale, scores are averaged with higher scores indicating higher amount of use or movement quality.

The SUS was adapted to capture the usability of wearable sensors.^33^ The adapted SUS consists of seven questions with five response options (Strongly agree to Strongly disagree) to capture complexity, ease of use, ease to learn, awkwardness and confidence in use. The scores for each question are added and transformed into a 0-100 scale (0=negative; 100=positive). To facilitate the interpretation of our data, mainstream wearable fitness devices (e.g., FitBit, Apple Watch, etc.) are rated between 61.4-67.6/100 on the SUS by neurotypical volunteers.^37^

#### 2.5.2. Wearable sensor measurement

MiGo captures time in active movement for each arm and arm use ratio (minutes of paretic arm activity/minutes of less-affected arm activity). In neurotypical adults, the mean (standard deviation) use ratio is 0.95(0.06), which indicates nearly equal durations of arm/hand movement during daily activity.^38^ Active movements were recorded in 15-min time bins across the monitoring period and were aggregated for each day. A custom software program was used to extract the raw data, and the maximum daily active time, for each sensor.

### 2.6. Data analysis

Descriptive statistics were used to summarize the data. Adherence was computed over the seven days of monitoring, even if some participants wore the sensors for a longer period. Wearing time was calculated from the 15-min raw data log. Wearing time was determined as the first time in the day when an increase in active time was noted within a 15-min bin to the last 15-min bin of active time at the end of the day for either sensor. Periods of inactivity during the day (e.g., during a nap) were not removed from the total wear time. To determine if wear time or paretic arm/hand use changed over time, repeated measures of variance were used. Hourly arm/hand use was also calculated and averaged across the wearing period to represent paretic and less-affected arm/hand activity. The coefficient of variation for each hour and each day was computed and averaged for the monitoring period.

Due to the small sample size, non-parametric statistics were used. Correlations between clinical measures (FMA-UE, CAHAI, Rating of Everyday Arm-use in the Community and Home, Motor Activity Log, Confidence in Arm and Hand Movement) and paretic arm/hand use were analyzed with Spearman’s rank correlation and 95% confidence intervals were computed with RStudio 2022.07.2+576. A significance value of p<0.05 was set for all statistical tests. Correlation coefficients between 0.70-1.00 were considered strong, 0.40-0.69 moderate and 0-0.39 weak.^39^ A cut-off score on the FMA-UE of 50/66 was used to classify the motor impairments severity levels, based on previous work demonstrating that stroke survivors with a score of >50 on the FMA-UE have significantly higher arm/hand use than those with a score <50.^40,41^ Recordings from the semi-structured interviews were transcribed verbatim and analyzed using thematic inductive analysis by two independent researchers.^42^ Any disagreements were resolved by discussion.

## 3. Results

### 3.1. Description of participants

A total of 32 participants were recruited but 2 did not meet the inclusion criteria (i.e., severe cognitive impairments and ambulation with assistance). Our final sample comprised 30 chronic stroke survivors. No drop out occurred during the monitoring period. The median FMA-UE score was 46.0 (range 18-66; see Table 1 for participant characteristics), with an equal split between participants <50 and >50 on the FMA-UE.

**Table 1.** Demographic and clinical characteristics of 30 participants with complete data.

| Characteristics | % or Median (IQR) |
| --- | --- |
| Gender |  |
| Men | 60.0% |
| Transman | 3.3% |
| Women | 36.7% |
| Age | 61.5 (48.5-64.9) |
| Race |  |
| Asian | 17.2% |
| Black | 17.2% |
| Native Hawaiian or Pacific Islander | 10.3% |
| White or Caucasian | 37.9% |
| More than one Race | 17.2% |
| Ethnicity |  |
| Hispanic | 30.0% |
| Non-Hispanic | 66.7% |
| Unknown/not reported | 3.3% |
| Time since stroke (years) | 7.3 (4.5-10.75) |
| Hemisphere affected by the stroke | Right, 41.4%<br>Left, 58.6% |
| NIHSS (/42) | 2.0 (1.0-3.0) |
| MoCA (/30) | 26.0 (22.5-27.0) |
| Edinburgh Handedness Inventory | Right: 93.3% |
| FMA-UE (/66) | 46.0 (25.0-59.0) |
| REACH (/6) | 2.0 (1.0-4.0) |
| MAL-14 |  |
| Amount of use (/5) | 2.5 (1.6-3.5) |
| Quality of Movement (/5) | 2.5 (1.4-2.4) |
| CAHAI-7 (/49) | 27.0 (10.8-43.8) |
| CAHM (/100) | 37.0 (21.0-72.0) |

**Abbreviations:** CAHAI-7: Chedoke Arm and Hand Activity Inventory - 7, CAHM: Confidence in Arm and Hand Movement, FMA-UE: Fugl-Meyer Assessment – Upper extremity, IQR: interquartile range, MAL-14:
Motor Activity Log 14, MoCA: Montreal Cognitive Assessment, NIHSS: National Institutes of Health Stroke Scale, REACH: Rating of Everyday Arm-use in the Community and Home.

### 3.2. Feasibility

#### a) Adherence and technical issues

87.6% of the monitoring period was valid. Out of 210 total days of data collection, 10 days were missing due to lack of adherence, 2 days for error with the server, and 14 days for sensors malfunction. One participant called the team to report sensor malfunction and eight follow-up phone calls were made after catching adherence or system issues. Reasons for lack of adherence were cold-like symptoms unrelated to the study (n=2), forget to charge or wear the sensors (n=3) or incompatible activities (n=2). Three participants chose not to wear the sensors for one or multiple days. Both participants with cold-like symptoms chose to add one day to the data collection to compensate for a missed day. Sensors’ average wear time/day was 12.6(0.2) hours. There was no significant change in the wear time (F=1.74, p=0.15) or the paretic arm use over days (F=1.41, p=0.24). The coefficients of variation were 0.20 and 0.62 for day-to-day and hour-to-hour variability within participants, respectively.

The main technical issues were errors with the server and sensor malfunction. Server errors occurred in five participants, but data were recovered for three of them, leading to two missing days in total. Sensor malfunctions were trouble synching with the gateway (n=3; 9 days) or broken sensor during the monitoring period (n=1; 5 days). Researchers walked participants through the procedures to resolve the synchronization issue, but two participants did not understand this procedure, even after demonstration and verbal guidance. Since feedback capability was disabled during the data monitoring period, some participants mentioned they did not know if the sensors were working properly or if data were being recorded.

> “I didn’t experience technical issues, but I wasn’t sure if it was working every day. I couldn’t tell because there’s no kind of feedback to me that meant it was working or it was on.”

#### b) Safety and comfort

Most participants did not experience issues related to safety or discomfort. In general, the wristbands were comfortable, and the elastic band was preferred to the silicone band. Three participants reported discomfort and difficulty to adjust the silicone band. One additional participant reported that the wristband interfered with his resting splint. Despite the use of an alternative wristband, challenges to don/doff the sensors were reported by those with severe motor impairments, with three receiving regular assistance from caregivers. Unrelated adverse events occurred in three participants: cold-like symptoms (n=2), fall that occurred after the monitoring period (n=1) and hospitalization due to low potassium levels (n=1).

> “I found the silicone band on the one wrist. I would get things caught in it. The elastic band was better.”

> “Putting them on was the hard part.”

#### c) Acceptance and satisfaction

The mean SUS was 85.4(13.0)/100, which indicates high usability. Most participants reported having a seamless experience with the data monitoring and forgot they had the sensors on. Many felt that wearing wrist sensors made them more aware of their behavior. Participants mentioned that knowledge of being monitored was a motivation to use their paretic arm and hand more.

> “Once it’s put on, I forget about it the rest of the day.”

> “It was wonderful. I didn’t mind one day doing it.”

> “Having to understand that [I was being monitored] kept me motivated. I am more aware of my movement. My affected side, I noticed it more so than last week.”

### 3.3. Associations between motor capacity and performance

The mean paretic arm use ratio was 0.50(0.19) and the hourly paretic arm use duration was 6.70(3.74) minutes during waking hours. Both were normally distributed. Paretic arm use ratio differed between participants with >50 FMA-UE (0.62(0.18)) and those with <50 FMA-UE (0.41(0.15), p=0.01). Measures of motor capacity both at the body function/structure and activity levels were positively correlated with the measure of performance captured by the wearable sensors (Table 2; Fig. 1 panels A and C). This suggests that **participants with** greater motor capacity **use** the paretic arm**/hand more**. The correlation between CAHAI and the wearable sensor data was the strongest (ρ = 0.713, p<0.001, Fig. 1B). Self-efficacy (Fig. 1D) was also moderately associated with paretic arm use ratio in the natural environment, with higher paretic arm/hand use in participants with higher confidence in their paretic arm/hand movements.

**Fig. 1.**
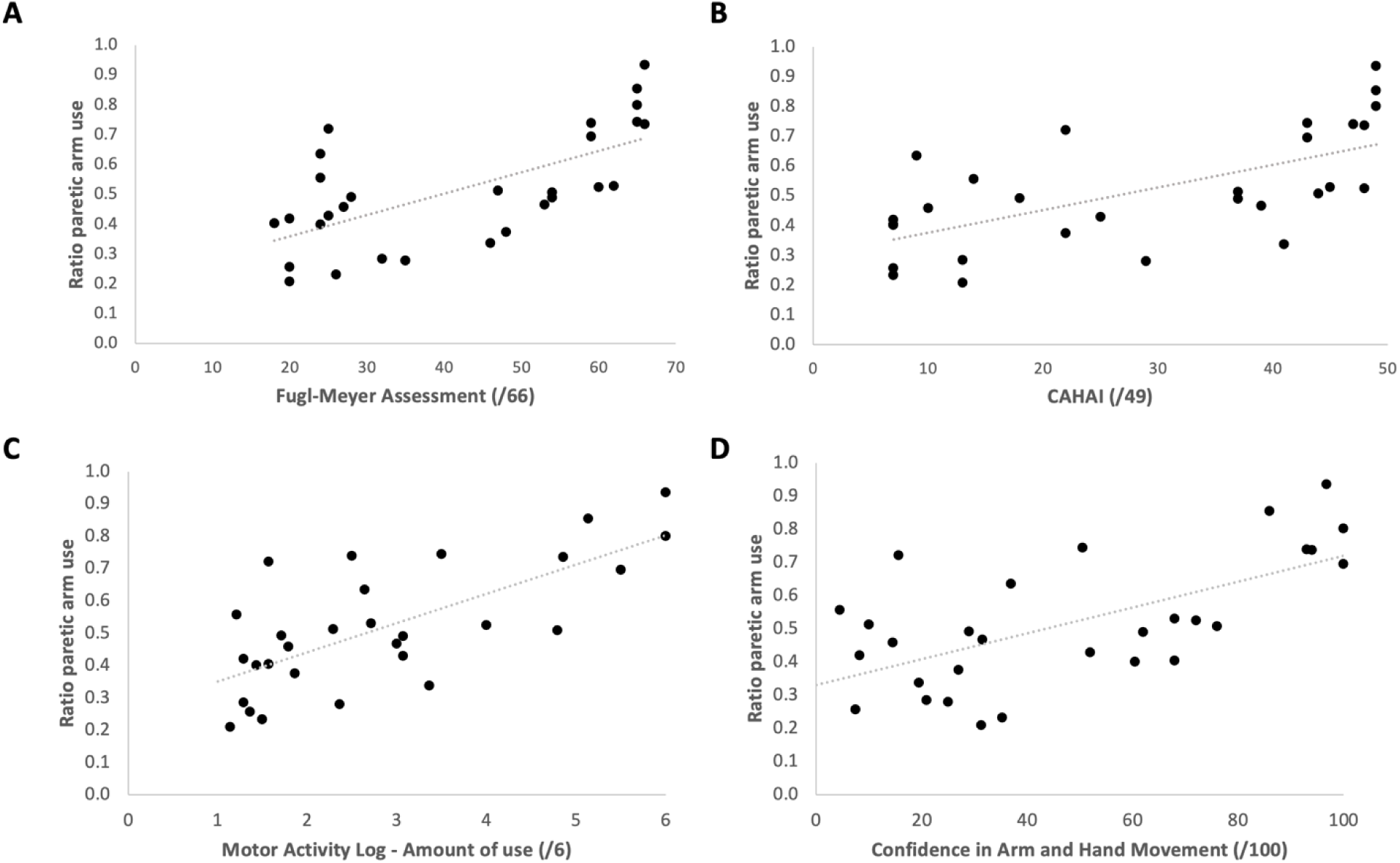
Relationship between wearable sensor data metrics and clinical assessment tools representative of specific domains of the International Classification of Functioning. A) Scatterplot of motor impairment (x-axis: Fugl-Meyer Assessment Upper-extremity score) vs. movement performance (y-axis: ratio of paretic arm use) in 30 chronic stroke survivors (Spearman correlation coefficient (ρ) = 0.67, p<0.001). B) Scatterplot of activity limitations in bimanual tasks (x-axis: Chedoke Arm and Hand Activity Inventory-7 score) vs. movement performance (y-axis: ratio of paretic arm use), (ρ = 0.71, p<0.001). C) Scatterplot of Motor Activity Log – Amount of use scale (x-axis) and ratio of paretic arm use (y-axis), (ρ = 0.66, p<0.001) between self-perceived paretic arm use and actual paretic arm use. D) Scatterplot of self-efficacy post-monitoring period (x-axis) and ratio of paretic arm use (y-axis), (ρ = 0.55, p = 0.01).

**Table 2.**
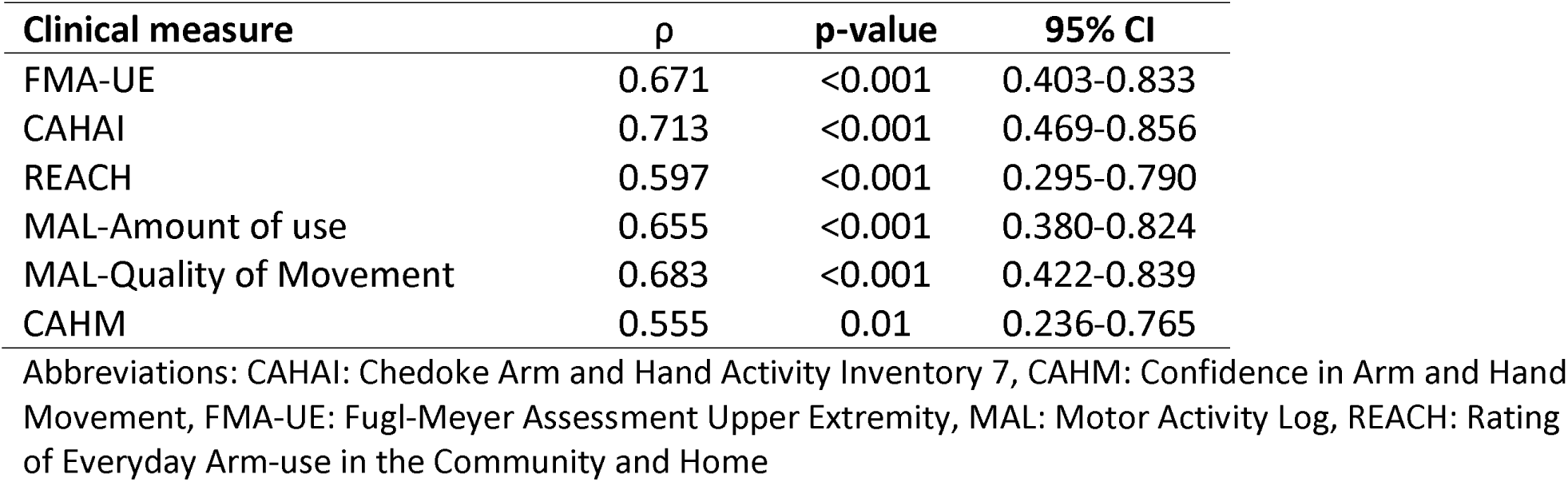
Spearman correlations (ρ) and 95% CI between paretic arm use ratio and arm motor impairments, functional capacity, perceived arm use, and self-efficacy.

There was a moderate positive correlation (ρ= 0.69, p<0.001) between the time in active movement of the paretic versus less-affected arm/hand. As less-affected arm/hand active movement increased, so did the paretic arm/hand active movement (Fig. 2A). Data visual inspection did not identify clear patterns of greater paretic arm/hand activity at certain times of the day between or within participants. Wearing schedule also varied between participants to accommodate their own schedule. On average, late mornings (10am-1pm) were periods of greater paretic arm/hand use with arm/hand activity slowly decreasing throughout the day (Fig. 2B) with large variability between participants. From the qualitative data, most participants reported that the monitoring period was representative of their typical activities. Nonetheless, the wearing schedule (12h/day) did not capture all the activities performed during a given day, as some participants made exercises in bed in the morning, walked their dogs or prepared breakfast before donning the sensors.

> “I decided I wasn’t going to do anything different, because I didn’t want to alter the data. I wasn’t going to pretend that I’m different.”

> “I didn’t put [the sensors] on until after I got dressed. Every morning I get up, eat breakfast and then I get dressed.”

**Fig. 2.**
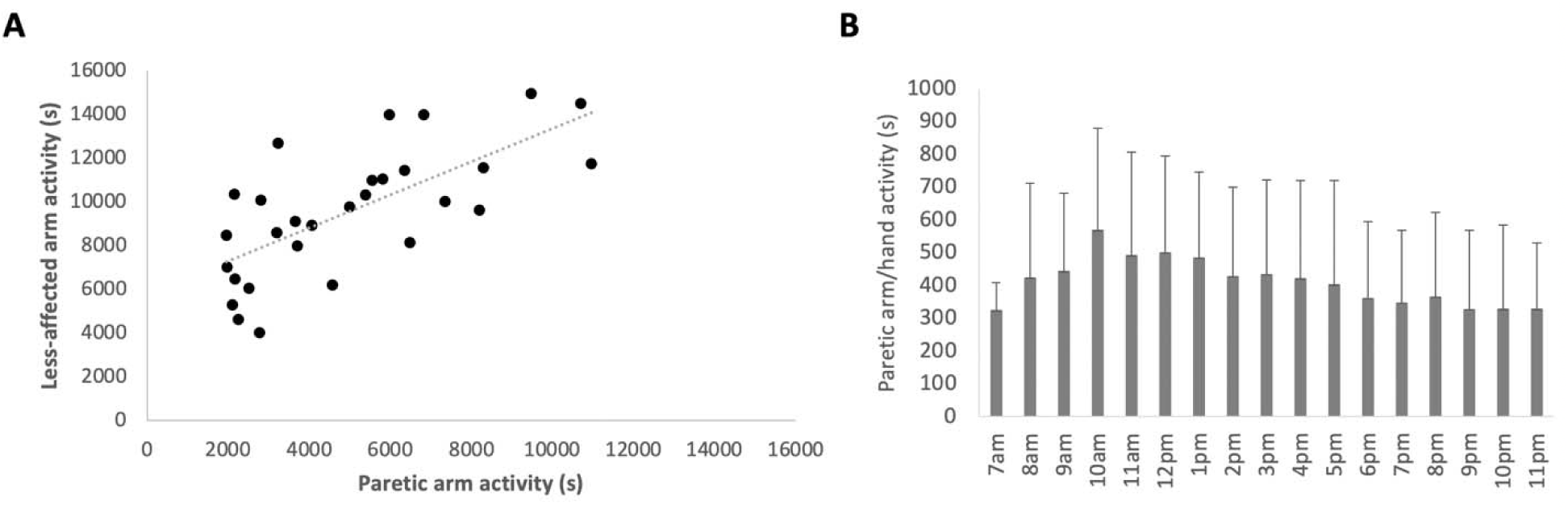
Scatterplot of sensor-derived paretic arm activity by Less-affected arm activity and paretic arm activity by hour across the seven days. A) Scatterplot of movement activity of the paretic (x-axis) and less-affected (y-axis) arms, showing a moderate positive relationship (ρ = 0.69, p<0.001). B) Average paretic arm/hand use (in seconds) for each hour of the day for 30 participants. Wearing schedule varies between participants, but all participants wore the sensors between 11am and 9pm. 4 participants donned the sensors at 7am, 15 at 8am, 22 at 9am, 28 at 10am. 1 participant doffed the sensors at 8pm, 4 at 10pm and 9 at 11pm.

## 4. Discussion

The results confirmed the short-term feasibility and usability of a consumer-grade wrist-worn sensor to capture arm/hand activity in chronic stroke survivors with mild-severe arm motor impairments, as all our milestones were met. Sensor-derived motor performance was closely related to clinical measures of motor capacity (impairments, activity limitations and self-perceived arm/hand use and quality) and self-efficacy. Our results are consistent with previous work done in the subacute stroke care setting demonstrating the relatedness between sensorimotor capacity and performance.^43–46^

In contrast to investigations of physical activity levels which require one week of data collection to capture inter-day variability,^47^ previous work examining post-stroke upper limb activity typically uses 24-72h monitoring periods.^15,38,40,43,48,49^ We chose a 12h wearing schedule to accommodate for MiGo’s battery life (∼72h) and the lack of waterproof enclosure. This wearing schedule can lead to the loss of important data, such as bathing and getting dressed. Water resistance and long battery life are important features to consider for future wearable technology. Based on the variability of the data, short data monitoring periods (24-72h) over 24h periods may be sufficient for research purposes. The fact that our participants rated a one-week monitoring period as acceptable bodes well for future clinical use where these longer monitoring periods may be necessary to better understand the multiple factors impacting arm/hand use after stroke in the natural environment.^35^ Of note, wear time did not decrease over the monitoring period, an indication that there were no apparent novelty or fatigue effects over the week.

The usability of the system was higher than SUS ratings from consumer-grade fitness devices.^37^ Nonetheless, discomfort with the silicone band and challenges to don/doff the sensors were raised. Consistent with our validity and usability findings,^20^ most participants were satisfied with the elastic band on the less-affected wrist. Elastic bands on both wrists should be considered for future use. For participants with severe motor impairments, individualized solutions to don/doff the sensors independently should be investigated. Options, such as a slap bracelet or a band that has open ends that curl around the wrist instead of fastening, could be explored, as those bands were preferred by stroke survivors and therapists as reported in a previous study.^50^ We took multiple steps to minimize missing data (e.g., tips sheet, daily monitoring of the web platform and follow-up phone calls). These efforts may have contributed to the high percentage of valid data but may be more difficult to accomplish in a clinical setting.

Some technical issues and malfunction occurred during this study, but this was expected at this stage of technology maturity. The use of a cellular gateway to transfer the data to a secured server allowed the research team to monitor adherence remotely and quickly identify and resolve technical issues. However, some sensors did not synch properly with the gateway and not all participants were able to learn the procedure to rectify the issue. Since MiGo feedback capabilities were disabled by design, participants could not identify if technical issues occurred. While it is not expected that feedback capabilities would be disabled when this technology is implemented outside the artificial research setting, this was highlighted as a limitation by our participants.

The results support our hypothesis that motor capacity and sensor-based performance measures are closely related, with the CAHAI having the strongest correlation. This is not surprising, as both measures captured bimanual arm use. Importantly, most daily activities require the use of both arms/hands. This phenomenon has been known for some time.^51^ The contribution of both arms/hands to daily activity is also supported by the close relationship between activity of the paretic and less-affected arms, which replicated the results from Bailey et al.^52^ However, while motor capacity and performance are related, our findings do not suggest that an improvement in motor capacity or self-efficacy will lead to an improved motor performance. Both Lang *et al*.^6^ and Doman *et al*.^53^ have demonstrated that improvements on standardized measures made after intensive rehabilitation do not translate to improvement in arm/hand use in the natural environment for most stroke survivors. Consistent with previous work,^40,41^ we found a significant difference between participants with a FMA-UE score <50 and those >50. This is aligned with the virtuous cycle of recovery hypothesis stating that for people with mild-moderate sensorimotor impairments, high levels of use and function reinforce each other.^12^ Recent work identified that stroke survivors can be categorized into five groups based on their arm/hand use performance measured with wearable technology. These groupings could be useful for clinical practice to guide clinical decision making and personalize care.^54^ The relationship between self-perceived arm/hand use and sensor-derived measures was higher than hypothesized. One possible interpretation based on our qualitative data is that the act of wearing the sensors, even with the feedback turned off, made participants more aware of their behavior, thus engaging participants in a more mindful evaluation of paretic arm/hand use in daily activities. Our results corroborate recent literature correlating self-efficacy and paretic arm use, and support self-efficacy as a factor that may explain the disparity between motor capacity and performance.^41,55–57^ Self-efficacy, or an individuals’ belief in their capacity to achieve certain outcomes, influences rehabilitation outcomes and, consistent with our findings, performance.^58^ We observed greater paretic arm/hand activity in the morning with activity slowly decreasing throughout the rest of the day. This might reflect a diminution in energy during the day, as poststroke fatigue is common.^59^ This work adds to the body of knowledge by providing promising implications for clinical practice: 1) encouraging stroke survivors to increase activity of the less-affected arm/hand may facilitate an increase in paretic arm/hand activity thereby leveraging the well-known prevalence of bimanual activities in the unsupervised setting, 2) interventions to enhance self-efficacy in therapy should be explored as a means to increase arm/hand use, and 3) mornings/early afternoons may be optimal time window targets for interventions.

### 4.1. Limitations

We recruited participants who chose to participate in our previous validity study and from an IRB approved database. As such, these volunteers may represent a group of early adopters of new technologies who are more compliant with study procedures than a typical chronic post-stroke population. The limitations of the wearable sensor data should be acknowledged in the interpretation of the relationship between capacity and performance, as wrist-worn wearable sensors cannot capture finger movements nor distinguish between purposeful and non-purposeful movements.^60^ It is possible that the administration of the Motor Activity Log after the monitoring period could inadvertently draw attention to their activity. **Finally, all** analyses were correlational, which prevents any causation conclusions to be drawn.

### 4.2. Conclusion

The feasibility of the commercial-grade wearable sensor system offers new possibilities for clinical practice to complement existing clinical assessments. Knowledge of spontaneous bimanual arm/hand use in the daily environment may provide a foundation for neurorehabilitation clinicians to 1) assess the transfer of skills gained in therapy to real-life situations, 2) guide personalized interventions and 3) evaluate progress.^50,61^ Participants’ perception of the usefulness of wearable sensors to encourage movement behavior supports the potential of wearable technology, not just as an assessment tool, but as a means to deliver real-time interventions outside the clinical setting. Future work should aim to develop theoretically driven and evidence-based interventions that leverage wearable technology to promote recovery-enabling behaviors.^12^

## Data Availability

All data produced in the present study are available upon reasonable request to the authors.

## 5. Acknowledgements

We acknowledge the contributions of Tanisha Gunby for assistance with data collection and data entry and of Courtney Koleda and Justine Buenaventura for their assistance with verbatim transcription and data verification.

