## Supplementary material for "Wearable technology to capture arm use of stroke survivors in home and community settings: feasibility and early insights on motor performance"

#### Tips sheet

**Equipment**

2 MiGo watches (right and left)
2 MiGo ankle sensors + bands (right and left)
2 MiGo bifurcated charging cables
1 Tenovi gateway
1 gateway charger

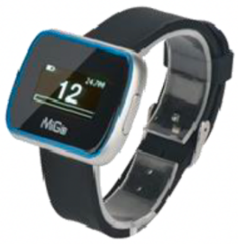


MiGo watch

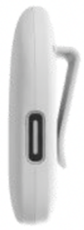

Ankle clip

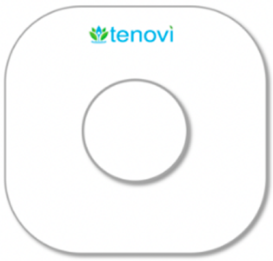


Tenovi gateway

**When to wear the MiGo:**

- Wear the MiGo watches, one on each wrist and the ankle clips, one strapped to each ankle, every day, during the entire week.
- Make sure the Left and Right sensors are on the appropriate Left/Right side of the body.

** The MiGo devices may feel a little strange at first but soon you will get used to them.

**How to charge the MiGo:**

A. Connect the provided power cord with the Tenovi gateway and the MiGo device chargers to an electrical outlet and keep the power cord plugged in at all times.
B. Keep the Tenovi gateway device connected to the plugged-in power cord at all times.
C. The MiGo devices (2 watches, 2 ankle clips) must be charged for at least 5 hours every night.
D. To plug in MiGo watches, gently open the charging cover (black), and attach to the charging cable. (See image)

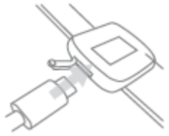

E. To plug in the MiGo ankle clips, attach to the charging cable.

**You should notice the MiGo devices 'beeping' shortly after you plug them to charge. This is normal!! It means they are uploading all your movements!

**Precautions**

- Do not wear the MiGo while charging it.
- MiGo is not water resistant. Remove all MiGo devices during showering, bathing, swimming or washing the dishes. Do not use the MiGo product in a sauna.
- If the MiGo devices get dirty, then use a damp towel to clean the MiGo devices. Do not use abrasive cleaners to clean your MiGo products.

### Interview guide

**Introduction:**
Thank you again for your participation in this study. We want to obtain your impressions and your satisfaction towards wearing the MiGo sensors for the past week. We also want to know your opinion about what influences you to use your most-affected arm in your daily activities and what influences you to move around your house or in the community. Before we start with the questions, I want to give you your Movement Report. We will go through this report together and I will ask you a few questions. This report provides information on your arm movements and your walking activity over the past week while you were wearing the MiGo.

During this interview, we would like to know what you think and how you feel about MiGo. You should try to say your honest and true opinion. Your feedback will help us to design an intervention that can be used in rehabilitation for stroke survivors like you. The discussion will take no more than 60 minutes. May I record the discussion to facilitate its recollection? *(if yes, switch on the recorder)*

**Confidentiality:**
Despite being taped, I would like to assure you that the discussion will be confidential. The recording will be transferred to a password-protected computer until it is transcribed word for word. Then, the recording will be destroyed.

1. **Overall experience (results from this section reported in this manuscript)**

- Do you think the previous week was representative of your typical everyday activities?
    - How was it different than usual?
    - Do you think wearing the MiGo sensors made you move more or less than what you would consider your 'normal' amount of activity?
- Please describe your experience wearing the MiGo sensors.
- How comfortable was each sensor?
- Tell me the problems you experienced during the study (i.e., technical issues, injury).
- What do you feel are some advantages of using the MiGo for rehabilitation?
- What do you feel are some disadvantages of using the MiGo for rehabilitation?

2. **Arm use in daily activities (results from this section reported elsewhere)**

- How much are you using your more affected arm compared to their less-affected arm**?**
    - How does it compare to before you had your stroke?
- Specifically, tell me some things that motivate you to use your more-affected arm to do your everyday activities?
- What limits you from trying to use your affected arm since your stroke?
- Please describe a few activities that are difficult for you to accomplish because of your arm.
- What do you think has influenced the recovery of your arm?

- <u>After</u> your rehabilitation stay, what (if anything) has most influenced how much you used your arm.

**3. Walking behavior in daily activities (results from this section reported elsewhere)**

- Thinking about the amount of walking activity you do ***inside your home***:
    - Do you feel you are more active, less active or about the same?
    - How do you feel about how well you're walking now as compared to before your stroke? Describe what you feel are some major differences with HOW you walk now as compared to before.
- Now thinking about the amount of walking activity you do ***outside of your home***:
    - Do you feel you are more active, less active or about the same?
    - How do you feel about how well you're walking now as compared to before your stroke? Describe what you feel are some major differences with HOW you walk now as compared to before.
- What factors or situations influence how much or how little you walk in your everyday activities?
- What would you say are the most limiting factors to you walking inside your home?
    - Outside your home?
- What do you do to overcome these problems or limitations with walking?
- What would motivate you to walk more?
- <u>After</u> your rehabilitation stay, what (if anything) has most influenced how much you walk in your everyday activities?
    - How well do you walk in your everyday activities?

**4. Perception of the Movement report (results from this section reported elsewhere)**

A. Let's look at the graphs that represent your arm movement.

- Tell me which graphs do you like the best?
    - Are there any graphs that are confusing to you?
- Which graph motivates you to move your affected arm more?

B. Now, let's look at the graphs that represent how much and how well you walk.

- Tell me which graphs do you like the best?
    - Are there any graphs that are confusing to you?
- Which graph motivates you to increase the amount you walk in your home?
    - In the community?
- Which graph motivates you to improve the quality of how you walk in your home?
    - in the community?

C. Feedback delivery

- How would you like the Movement report to be offered? (e.g., on an app on your phone, by email, on the watch itself, during a meeting with a clinician)
- How would you use this Movement report?
    - If feedback was offered on the watch or on an app on your phone, would you use it?

- How useful do you find this type of feedback to increase/improve the movements of your arm/walking?

D. How can the Movement Report be improved?

E. Is it helpful to receive objective feedback (like the Movement Report) on your activity?
   - How often would you prefer to receive this feedback?
     - More than once per day, daily, weekly, or less than once per week?
   - Now that we have discussed the Movement Report, do you feel that you could navigate and use this information on your own (or with help from a caregiver)?
   - How would you prefer to receive feedback? (e.g., delivered through the Movement Report? by speaking with someone directly? a combination of both? or a different method?)

**Summary of the discussion**

Let's summarize some of the key points from our discussion.

- Is there anything else you would like to add?
- Do you have any questions?

**Conclusion**

Thank you for participating in this study. This has been a very successful discussion.
Your opinions will be an asset to this study.
